# Natural Language Processing Algorithms Outperform ICD Codes in the Development of Fall Injuries Registry

**DOI:** 10.1101/2024.09.26.24314444

**Authors:** Atta Taseh, Souri Sasanfar, Jia-Zhen M. Chan, Evan Sirls, Ara Nazarian, Kayhan Batmanghelich, Jonathan F. Bean, Soheil Ashkani-Esfahani

## Abstract

**Background:** Standardized registries are commonly built using administrative codes assigned to patient encounters, such as the International Classification of Diseases (ICD) codes. However, fall patients are often coded using subsequent injury codes, such as hip fractures. This necessitates manual screening to ensure the accuracy of data registries. Herein, we aimed to automate the extraction of fall incidents and mechanisms using Natural Language Processing (NLP) and compare this approach with the ICD method.

**Methods:** Clinical notes for patients with fall-induced hip fractures were retrospectively reviewed by medical experts. Fall incidences were detected, annotated, and classified among patients who had fall-induced hip fracture (case group). The control group included patients with hip fracture without any evidence of fall. NLP models were developed using the annotated notes of the study groups to fulfill two separate tasks: fall occurrence detection and fall mechanism classification. The performances of the models were compared using accuracy, sensitivity, specificity, positive predictive value (PPV), negative predictive value (NPV), F1-score, and area under the ROC curve (AUC-ROC).

**Results:** A total of 1,769 clinical notes were included in the final analysis for the fall occurrence task, and 783 clinical notes were analyzed for the fall mechanism classification task. The highest F1 score using NLP for fall occurrence was 0.97 (specificity=0.96; sensitivity=0.97) and for fall mechanism classification was 0.61 (specificity=0.56; sensitivity=0.62). NLP could detect up to 98% of the fall occurrences and 65% of the fall mechanisms accurately compared to 26% and 12%, respectively, by ICD codes.

**Conclusion:** Our findings showed promising performance with a higher accuracy of NLP algorithms compared to the conventional method for detecting fall occurrence and mechanism in developing disease registries using clinical notes. Our approach can be introduced to other registries that are based on large data and are in need for accurate annotation and classification.

## 1. Introduction

With 3 million emergency room visits, 300,000 hospitalizations, and 30,000 fatalities annually, falls pose a major threat to public health.^1,2^ The financial impact is also substantial, with an estimated $50 billion in medical expenses for non-fatal falls.^3^ Therefore, researching and understanding the nature of falls and fall-related injuries are crucial for developing effective prevention and treatment strategies as the populations age.^4^ Given the multifactorial nature of falls, and the difficulties of conducting prospective research in the field, developing fall registries comprised of large, and accurate medical data, is of great importance.^5,6^ Standardized registries are commonly built using administrative codes, such as the International Classification of Diseases (ICD), that are assigned to patient encounters, and Current Procedural Terminology (CPT) codes. Previous studies have therefore used these codes to extract patients with a history of falls ^9–11^. However, this method has limitations that may lead to an underestimation of actual fall frequency and might not reveal the history of falls in patients.^12^ Reporting falls using the External Causes of Morbidity codes is usually recommended but not mandatory in all healthcare settings. Since falls are not typically considered standalone conditions, many healthcare providers may rather use the diagnosis ICD codes and assign codes to the end result of a fall – e.g. a hip fracture, rather than the fall itself.^13,14^ This makes it difficult for investigators to identify falls in medical history of the patient and the true frequency of falls within populations. Given these limitations, clinical notes were suggested as a more reliable methods of detecting falls, fall mechanisms, and fall-induced injuries.^8^ This process, however, is expert dependent and time-consuming, particularly if the dataset is large. To address these obstacles, natural language processing (NLP), which combines computational linguistics and deep learning models to process narrative data, can be utilized to automate the review process of clinical notes to detect falls.^8^

This study aimed to assess the performance of NLP algorithms compared to the conventional methods of detecting fall incidence, and mechanism of falls obtained from clinical notes of patients with hip fractures. Our hypothesis is that NLP algorithms outperform ICD codes in the detection of falls and fall mechanisms in patients with hip fractures.

## 2. Materials and Method

### 2.1 Study Design and Data Sources

A retrospective case-control study was conducted under the IRB number 2023P000741 at four tertiary hospitals located in Boston area, Massachusetts. Data was retrieved through the institution’s data repository using CPT codes for hip fractures (27125, 27130, 27226, 27228, 27235, 27236, 27244, 27245 and 27248) between January 2010 and December 2019.

Electronic health records (EHR) were screened for the presence of falls where a fall was defined as “an unintentional event that results in the person coming to rest on the ground or another lower level”.^17^ Patients ≥18 years old with a history of hip fracture were obtained and their medical notes were screened. Based on the notes, the patients were categorized into cases (who had falls and a hip fracture after the fall) and controls (who had hip fractures not due to falls). Falls resulting from violent encounters, animal attacks, significant external forces such as car or motor vehicle accidents, high-impact sports like skiing, and fractures caused by underlying pathological conditions were excluded to reduce the heterogeneity of fall mechanics. This exclusion helps avoid the influence of confounding injuries that differ significantly from typical accidental falls, ensuring that the study focuses on more clinically relevant fall types (Figure 1.). We included single notes for each patient in the case group since falls are directly documented alongside fractures. In contrast, multiple notes were reviewed and included for controls to ensure fractures are not associated with falls, providing a more comprehensive review of clinical history. The mechanisms of fall (the way falls happened) were further classified into three categories - same-level (S; occurring on the same plane or surface), multi-level (M; descent from one level to a different one), and unclassified (U; not classifiable due to lack of sufficient information).^18^ The annotations were done by an experienced orthopaedic scientist (AT) and the decisions for equivocal or debatable cases were made by a senior scientist (SAE). Expert annotations, serving as the ground truth for training the NLP models, were derived directly from clinical notes. Therefore, discrepancies between the documented fall mechanisms in these notes and the corresponding ICD codes compromised the validity of comparisons between ICD and NLP-based approaches. Consequently, patients with conflicting information between clinical notes and ICD codes regarding the fall mechanism were excluded to ensure the integrity of the analysis (Figure 1.).

**Figure 1.**
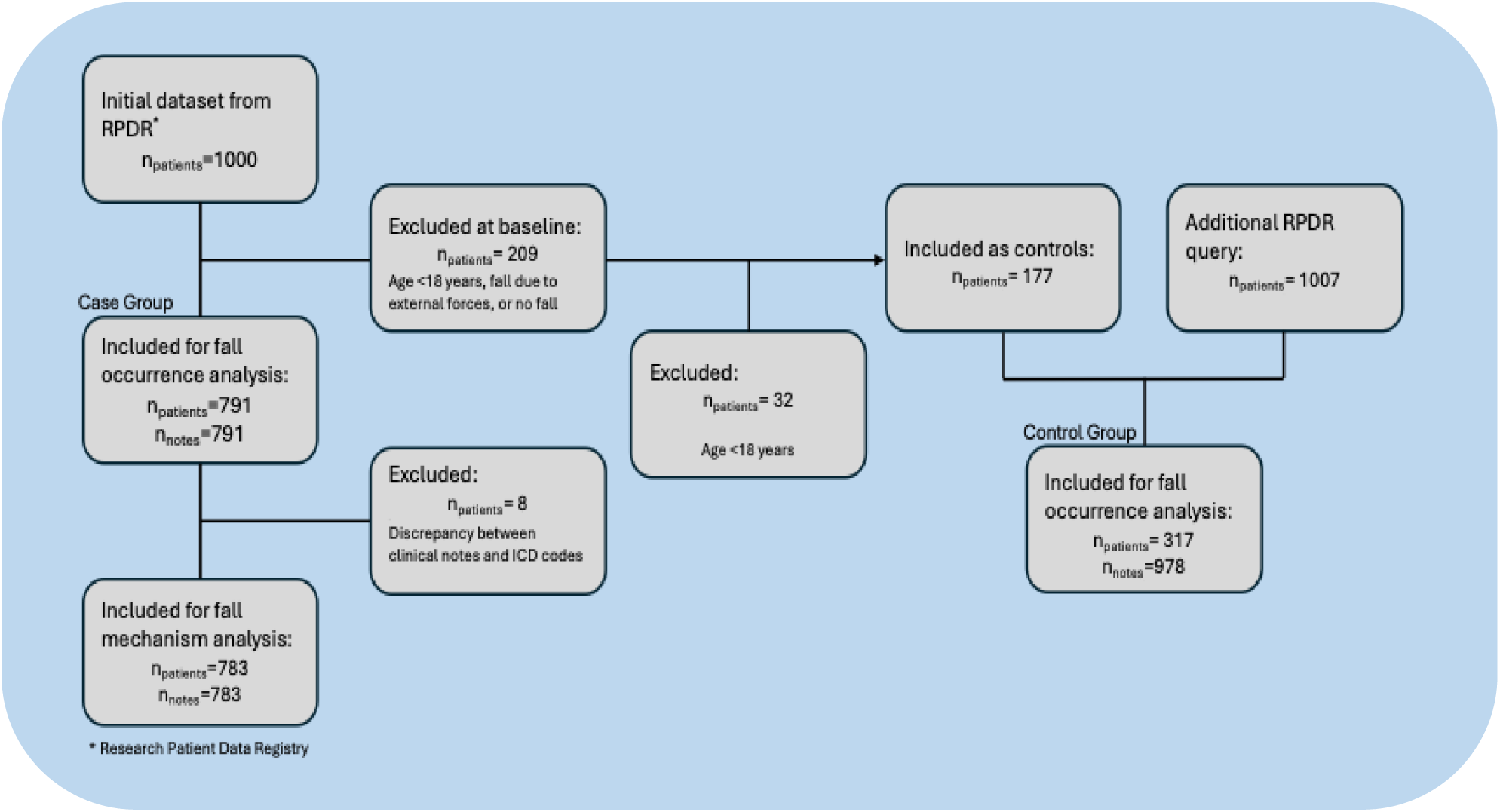
Study population flowchart

### 2.2 Data Preprocessing

A variety of unstructured clinical notes including history and physical examination, discharge summary, progress, operation, and emergency department notes were obtained. Due to the diverse formatting of these clinical notes, specialized preprocessing methodologies were required, which diverged significantly from conventional text processing approaches. Following annotation, the clinical notes underwent various preprocessing steps including de-identification, segmentation, and cleaning.^19^ The specific techniques used in preprocessing, which address the unique challenges posed by the clinical notes’ formatting, are outlined in Table 2. Detailed information about the segmentation process is provided in Appendix A. This detailed account ensures that the data is optimally prepared for the subsequent analytical phases.

### 2.3 Model Development

Models were developed to automate two distinct tasks including fall occurrence and fall mechanism classification. For the binary task of fall occurrence (fall vs no fall), a data split of 80:20 was used for training and testing purposes, respectively. Our methodology harnessed the text analysis capabilities of a modified Bidirectional Encoder Representations from Transformers (BERT) model described by Fu et al.^15^ We employed a maximum sequence length of 512 tokens, consistent with the recommendations in the original study by Devlin et al., used a batch size of 8, and conducted training over 3 epochs. Moreover, the Adaptive Boosting (AdaBoost) algorithm was utilized for fall identification, using single-layer decision trees (stumps) as described by Quinlan et al.^20,21^ AdaBoost assigns coefficients based on each classifier’s performance and adjusts sample weights during training to emphasize previously misclassified samples. Our hybrid classifier integrated Term Frequency-Inverse Document Frequency (TF-IDF) vectorization of textual data with AdaBoost, designed to handle weighted inputs and meet the study’s textual analysis requirements. The AdaBoost classifier was configured with 200 estimators, a learning rate of 1, and a base estimator of a decision tree classifier with a maximum depth of 5, using the SAMME algorithm. Lastly, Extreme Gradient Boosting (XGBoost), which is a refined version of gradient boosting recognized for its precision and versatility. XGBoost is noted for its use of sequentially connected classifiers, where each classifier builds upon the residuals left by the previous one. The XGBoost classifier was configured with a maximum depth of 3, a learning rate of 0.2, 200 estimators, verbosity set to 1, colsample_bytree of 0.5, and the evaluation metric set to ‘mlogloss’. ^22^

To address the challenges posed by the complex multi-class scenario in the fall mechanism classification task, which involved detailed classification into three categories (S, M, and U classes), we designated 70% of the data for training and 30% for testing. We employed a comprehensive suite of advanced machine learning models including AdaBoost, Support Vector Machine (SVM), XGBoost, and Random Forest (RF). Each model was chosen for its proven ability to decipher complex data relationships and offer detailed insights into the correlated factors of falls across the varied categories.^23–28^ The SVM model, a sophisticated two-layer recognition method, excels in identifying linear classifiers that maximize the separation distance within a dataset’s feature space.^29^ The model was configured with probability set to True, a regularization parameter (C) of 10, a radial basis function (RBF) kernel, a degree of 3, and gamma set to ‘scale’. RF, an ensemble learning method that constructs multiple decision trees during training and merges their results to improve predictive accuracy and control overfitting, was configured with 200 estimators, a maximum depth of 30, a minimum of one sample per leaf, and a minimum of 10 samples required to split an internal node.^30^ The XGBoost classifier was configured with an objective of ‘multi’, a maximum depth of 5, a learning rate of 0.3, 100 estimators, and the evaluation metric set to ‘mlogloss’. Finally, the AdaBoost classifier was configured with 200 estimators, a learning rate of 1, and a base estimator of a decision tree classifier with a maximum depth of 5, using the SAMME algorithm.

### 2.4 Statistical Method

Comparison of the baseline characteristics was done using SPSS software (IBM SPSS Statistics, Version 28), where T test and Chi square were utilized for continuous and categorical data, respectively. Several metrics were employed to evaluate the models’ performance in identifying and classifying falls. These metrics included sensitivity, specificity, F1 Score, positive predictive value (PPV), negative predictive value (NPV), accuracy, and area under the receiver operating characteristics curve (AUC-ROC). For multi-class classifications a weighted-averaging approach was used to report the overall model performance.^31^ Furthermore, the percentage of the notes correctly classified for each task by machine learning and ICD approach were calculated and compared by means of Chi-square testing. A 0.05 type one error probability was considered significant.

## 3. Results

A total of 1,769 clinical notes were analyzed for the fall occurrence task. Of these, 791 notes corresponded to the case group (one note per patient, n=791), and 978 notes were from the control group (representing 317 individuals with multiple notes per individual). Moreover, for the fall mechanism classification task, 783 notes (one note per patient, n=783) were included comprising 511 same-level falls, 151 multi-level falls, and 121 unclassified falls. The case group comprised of older individuals with a mean age of 77.7 ± 14.3 years versus 65.3 ± 19.6 years of the control group (p<0.001; Table 1). Furthermore, although both groups had a higher proportion of females, the case group had a notably higher percentage of female patients compared to the control group (p=0.01, Table 1).

**Table 1.**
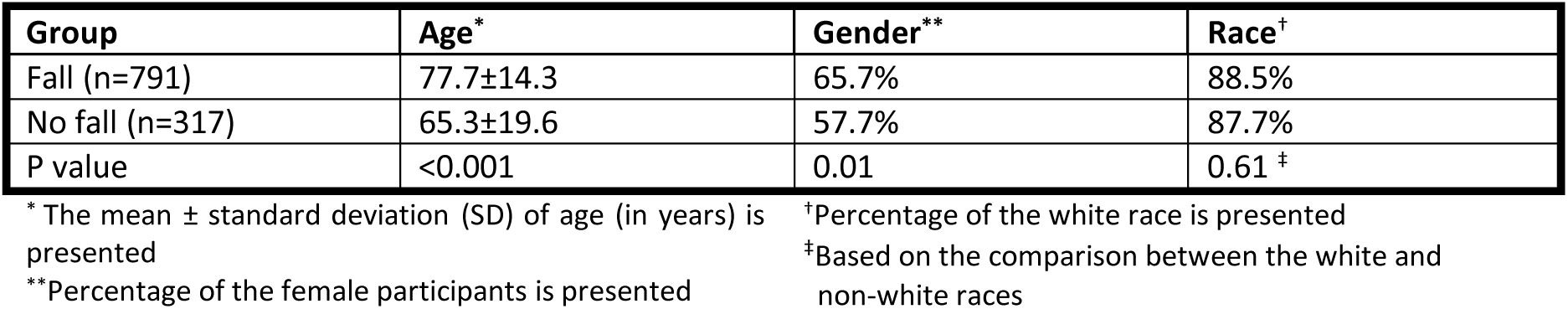
Comparison of the baseline characteristics of the study groups.

| Group | Age* | Gender** | Race† |
| --- | --- | --- | --- |
| Fall (n=791) | 77.7±14.3 | 65.7% | 88.5% |
| No fall (n=317) | 65.3±19.6 | 57.7% | 87.7% |
| P value | <0.001 | 0.01 | 0.61 ‡ |

\* The mean ± standard deviation (SD) of age (in years) is presented
\*\*Percentage of the female participants is presented
†Percentage of the white race is presented
‡Based on the comparison between the white and non-white races

**Table 2.** An overview of the data preprocessing stages.

| Stages | Tool/Method | Purpose | Output |
| --- | --- | --- | --- |
| De-Identification | Stanford de-identifier | Remove personal identifiers to ensure privacy and compliance with data protection regulations. This involves replacing all PHI entities with synthetic variants to maintain data integrity and eliminate biases. The model chosen was the Stanford-de-identifier-base-model developed by Chambon et al, with an F1 score of 98.9 on the I2b2 2014 test set.<sup>41</sup> | Anonymized text ready for analysis. |
| Segmentation | Bespoke parser, FSM, and regular expressions | Segment notes into distinct sections for enhanced text processing accuracy. The parser identifies section headings and concatenates segments, refined through manual evaluation and iterative improvements. More details are provided in Appendix A. | Accurately segmented text with sections tagged for reassembly. |
| Filtering Uninformative Data | Identification and removal:<br>- Duplicates<br>- Uninformative sections<br>- Administrative content | - Remove duplicated sections from notes to prevent skewing results.<br>- Discard sections containing only headings without informative text.<br>- Remove document finalization and signature sections marked with terms like “signed” and “FINAL.” | Dataset free of redundant and uninformative sections. |
| Elimination of Non-Essential Elements | Regular expressions and manual filtering | Exclude conversion error notifications, Unicode/hexadecimal sections, and other irrelevant elements. | Dataset without non-contributory headers, unreadable sections, and irregular patterns. |
| Removal of Irrelevant Metadata | Manual filtering | Remove timestamps, de-identified placeholders, and other non-analytical metadata. | Dataset without timestamps and placeholder text, ensuring grammatical consistency. |
| Splitting the Data | Random allocation | Partition the dataset into training and testing subsets for unbiased model evaluation. | Training and testing subsets for model development and performance evaluation. |

For detecting fall occurrences, all three models performed well, with the BERT model showing a lower F1 score and AUC-ROC (Table 3, Figure 2). The models could successfully classify a significant portion of patient notes (XGBoost=97%, AdaBoost=98%) as opposed to the ICD approach which could find 26% of them (p<0.001; Table 4).

**Figure 2.**
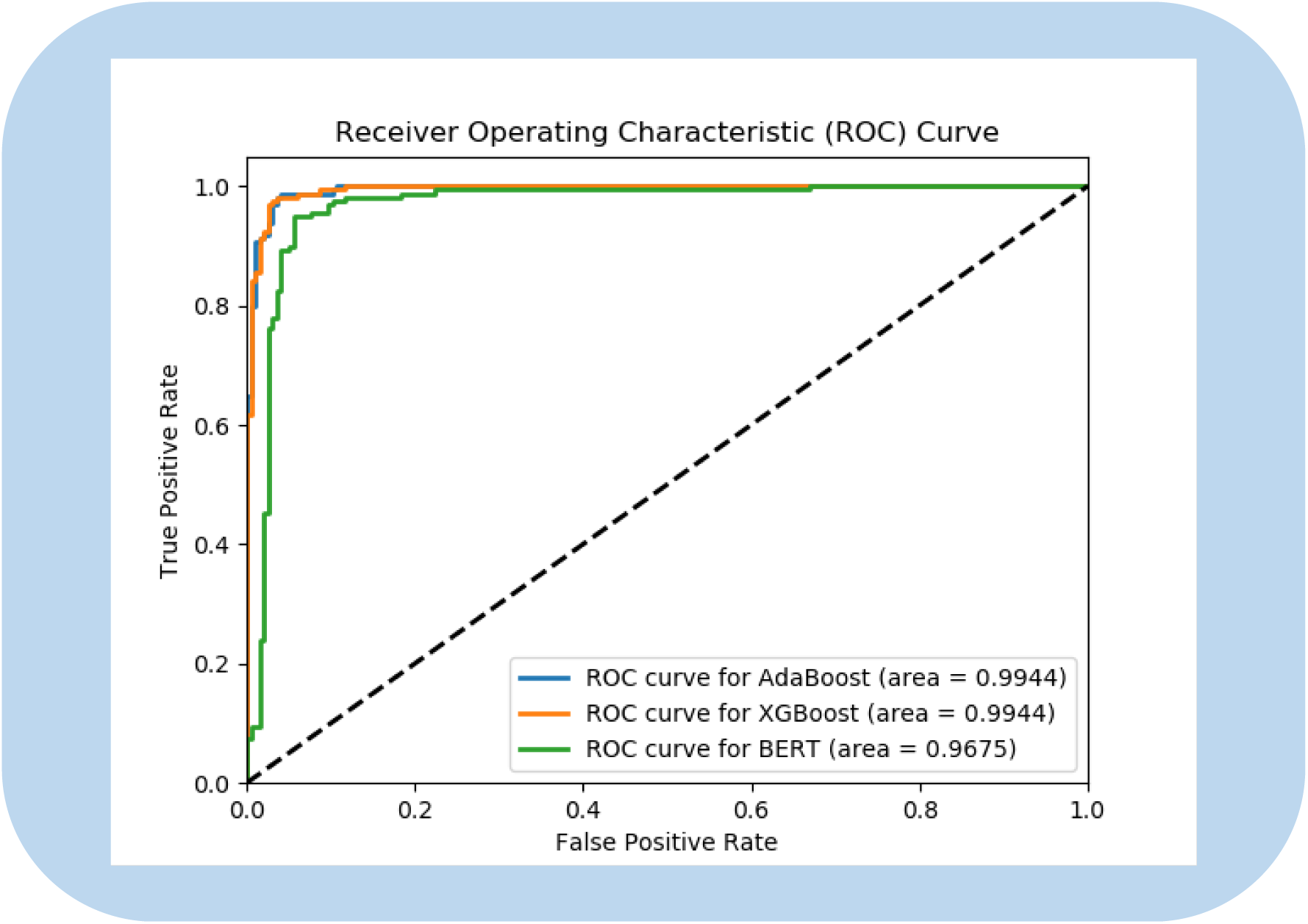
Receiver Operating Characteristics Curve (ROC) for the fall occurrence detection task

**Table 3.** The performance metrics of the study models for detection of fall occurrence and fall mechanism classification. Algorithms were trained on an expert annotated database.

| Outcome | Model | PPV | NPV | Sensitivity | Specificity | F1-Score | Accuracy | AUC-ROC |
| --- | --- | --- | --- | --- | --- | --- | --- | --- |
| Fall Occurrence Detection | BERT | 0.94 | 0.88 | 0.84 | 0.96 | 0.88 | 0.90 | 0.97 |
|  | AdaBoost | 0.95 | 0.98 | 0.98 | 0.96 | 0.97 | 0.97 | 0.99 |
|  | XGBoost | 0.96 | 0.98 | 0.97 | 0.96 | 0.97 | 0.97 | 0.99 |
| Fall Mechanism Classification* | SVM | 0.56 | 0.50 | 0.62 | 0.36 | 0.57 | 0.62 | 0.67 |
|  | AdaBoost | 0.55 | 0.43 | 0.60 | 0.39 | 0.56 | 0.60 | 0.61 |
|  | XGBoost | 0.60 | 0.51 | 0.62 | 0.56 | 0.61 | 0.62 | 0.65 |
|  | RF | 0.60 | 0.52 | 0.65 | 0.35 | 0.60 | 0.65 | 0.70 |

\*Weighted metrics are presented
Abbreviations: PPV, Positive Predictive Value; NPV, Negative Predictive Value; AUC-ROC, Area Under the Receiver Operating Characteristics Curve; RF, Random Forest; SVM, Support Vector Machine; BERT, Bidirectional Encoder Representations from Transformers; AdaBoost, Adaptive Boosting; XGBoost, Extreme Gradient Boosting

**Table 4.**
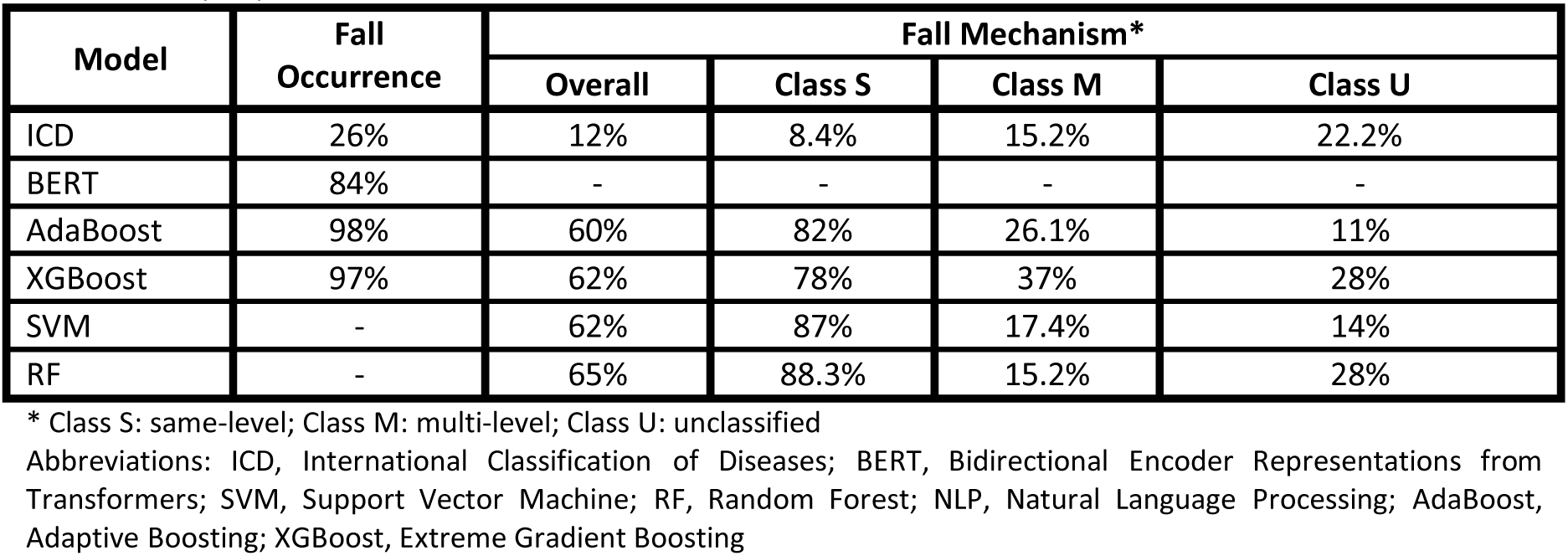
Percentage of fall notes correctly classified by NLP using clinical notes versus obtaining the notes using ICD codes. Data included notes of patients who had hip fractures with or without fall injuries, annotated by experts.

Regarding fall mechanism classification, the RF model slightly outperformed the others with an AUC-ROC of 0.70 and an F1 score of 0.60 (Table 3, Figure 3). Moreover, the RF model was able to correctly classify fall mechanism in 65% of the fall notes compared to the 12% of the ICD method (p<0.001, Table 4.). However, all four NLP models showed high classification performance in identifying class S falls only (Table 4).

**Figure 3.**
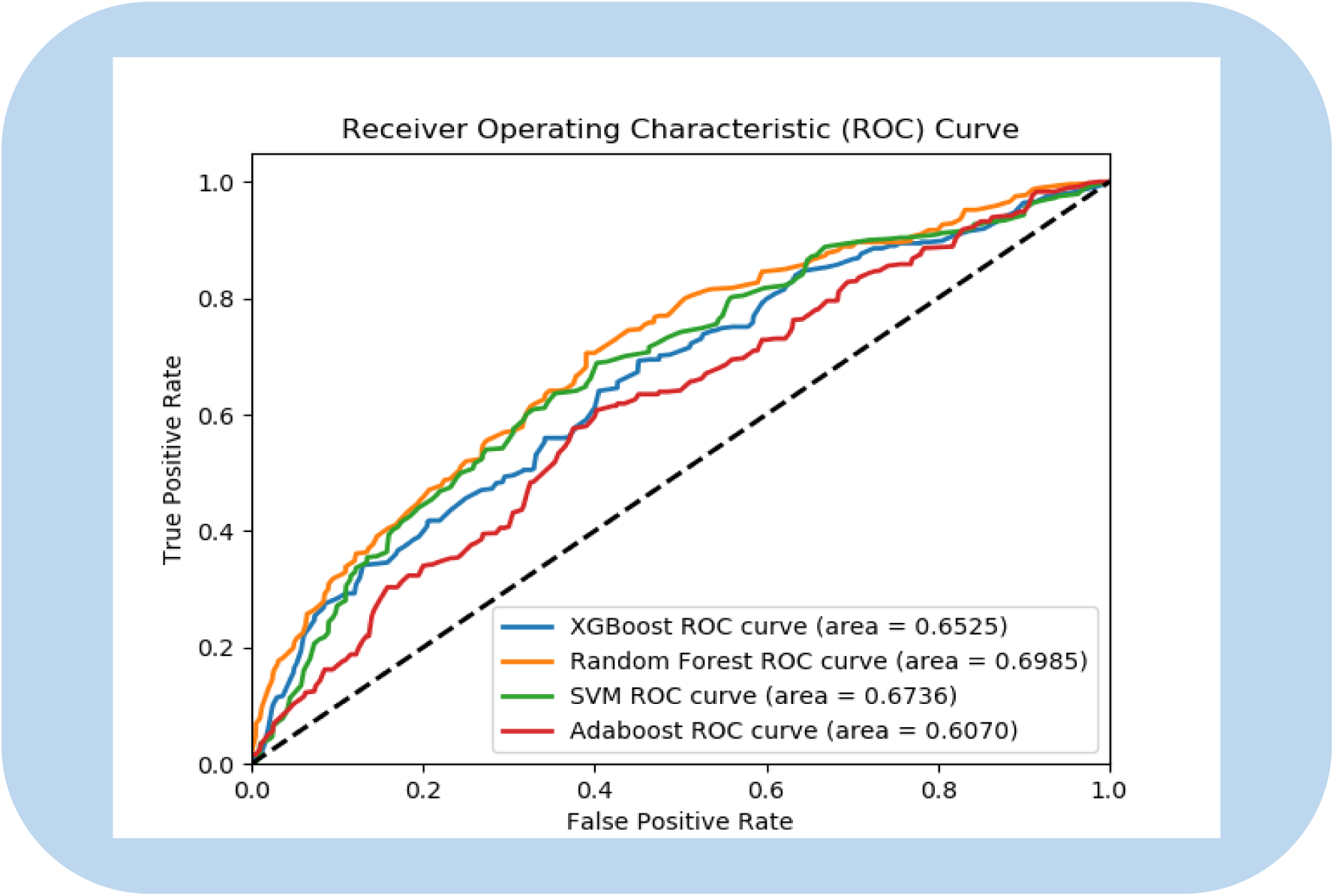
Receiver Operating Characteristics Curve (ROC) for the fall mechanism classification task

## 4. Discussion

This study aimed to automate fall identification and classification based on its mechanism from clinical notes and subsequently compare the results with the traditional ICD approach for building fall registries. Our results demonstrated superior performance of NLP models, which correctly identified 98% of the notes for fall occurrence compared to the 26% detected by the ICD approach. Furthermore, the models were able to classify 65% of fall mechanisms while the ICD approach detected 12% of these cases. Automated identification of fall incidents from clinical notes is an emerging topic in the realm of biomedical sciences. It serves multiple purposes such as insurance claim processing, cost analysis for falls, and enhancing fall prevention measures for inpatient safety, among others.^32–34^ Despite these varied objectives, there are commonalities in the methodologies and models employed. However, the interpretation of results can vary significantly and must be tailored to the specific study goals. Cheligeer et al. highlighted the superior performance of BERT and machine learning models in detecting inpatient falls compared to traditional ICD coding.^35^ Their findings underscored these models’ ability to accurately identify non-fall cases, as evidenced by high NPV and specificity. Nevertheless, when aiming to develop a comprehensive registry, achieving optimal sensitivity to maximize the inclusion of fall patients, alongside a high F1 score to balance PPV and sensitivity, become crucial.

Classical machine learning methods are commonly employed in fall classification studies. Luther et al. developed an SVM model using free-text clinical notes and a term-document matrix for feature selection, achieving an F1 score of 0.87.^36^ Our study extends this by employing a TF-IDF feature selection method, which weighs terms based on their importance to capture nuanced information from the notes. We found that ensemble methods achieved optimal performance with an F1 score of up to 0.98. Santos et al., have demonstrated superior performance of neural networks over classical machine learning methods.^37^ This finding is supported by Fu et al., who showed high performance of context-aware models like BERT in fall detection tasks.^15^ However, in our study, BERT did not outperform other machine learning models. BERT’s effectiveness is known to depend on the availability of sufficient training data due to its deep learning architecture.^38^ Therefore, the sample size in our study may have influenced the effectiveness of training within this framework.

Identifying fall mechanisms from patient records presents a significant challenge which if addressed properly can provide invaluable information for clinical and quality improvement purposes. Roudsari et al. investigated the acute cost of care for falls in patients over 65 years old, categorized by ICD codes for mechanisms.^14^ They found that same-level falls were the most common mechanism of injury (28%). However, the majority of falls (60%) were coded as an unspecified fall without a mention of the mechanism. In our study, only 11% of the notes were coded specifically for falls, and surprisingly, there were occasional discrepancies between the coded mechanisms and those described in clinical notes. Whether this discrepancy stems from insufficient clinical information or a tendency among providers to prioritize documenting immediate medical needs requires further investigation. Relying solely on medical coding seems not to be a reliable approach for identifying fall mechanisms.

While NLP has shown promise in retrieving data from medical records, its application in fall mechanism extraction remains underexplored. Liu et al. automated the extraction of inpatient fall severity from incident reports, leveraging structured features to improve the F1 score by 8%, achieving 0.78.^39^ In our study, we incorporated diverse types of unstructured clinical notes, including discharge summaries and progress notes. These notes were authored by various medical professionals with differing styles and descriptions of falls, introducing significant variability that posed challenges for extracting features. Our results indicated that the XGBoost and RF models achieved the highest F1 scores (0.6). These findings are consistent with previous research demonstrating improved disease classification accuracy using ensemble methods applied to medical notes.^39^ Additionally, using ensemble methods, Albano et al. have shown promise in enhancing the classification accuracy when dealing with rare classes.^40^ However, our study revealed suboptimal performance of the models in managing the ‘M’ and ‘U’ subclasses, likely due to the overall limited number of notes available for these classes. We suggest future studies should focus on increasing the dataset size for these subclasses to improve model performance.

Although this study was pioneering in addressing the development of fall registries, it had a few limitations. The sample size in our study seems sufficient for retrospective studies; however, for a machine learning study and in order to train the models appropriately, larger and more granular populations are needed. There was an imbalance in the S and M classes of the fall mechanisms, which significantly impacted the performance of the models. Although the class imbalance was a true representation of the real-world situation where most falls in the older adult population occur from standing, this could have limited the performance outcomes of our NLP models.

## 5. Conclusion

Our findings demonstrated a promising performance of NLP methods in identifying patients with a history of falls and hip fractures and their fall mechanism from clinical notes. This approach can significantly enhance the accuracy and efficiency of developing fall registries. Moreover, the models were particularly effective in classifying the mechanisms of falls in patients who experienced same-level falls. Future studies with larger sample sizes and a broader spectrum of pathologies can further validate these findings and address the issue of class imbalance. If well-expanded and developed, our approach can be introduced to the healthcare systems as an efficient and cost-effective approach for developing valid and reliable registry systems of diseases or clinical conditions that impose a great burden on the healthcare systems and the patients.

## Data Availability

All data produced in the present study are available upon reasonable request to the authors and approval of the institution's IRB

## Acknowledgment

We gratefully acknowledge the patients whose clinical data served as the foundation for this research, enabling us to advance the field of automated fall detection.

