## Appendix A for "Natural Language Processing Algorithms Outperform ICD Codes in the Development of Fall Injuries Registry"

**Supplementary Table 1.** Overview of the text segmentation process

| Condition | Sub condition 1 | Sub condition 2 | Example |
|---|---|---|---|
| Contains a word followed by colon | Segment contains no period in first 10 characters | First letter is capitalized | “Admission date:” |
| | Contains a ‘.’ in the first 10 characters.<br><br>(indicates a trail belonging to the previous section) | Part after ‘.’ should contain at least 1 character | “and the admitting diagnosis is shoulder pain. Check-up date: Saturday.”<br><br>(The text before the first period is added to the previous section, while the content after becomes a new section heading. |
| Consists of all uppercase character | First character is not a digit<br><br>(Exclude time-related information “7 AM”, dosages “500 MG PO”, and prescription list “1. VITAMIN D3” | | “HISTORY & PRESENT ILLNESS” |

**Supplementary Table 2.** This algorithm parses a block of text, segments it into sections based on predefined indicators, and stores these sections in a structured format.

```
Initialize an empty list to store the final list of complete sections.
Initialize the section head as None.
Split the text into parts based on newline characters ('\n').
Remove trailing white spaces and empty segments.
for each segment obtained from the split do
    if segment contains indicators for being section head, then
        if segment meets full conditions to be section head, then
            Make segment section head
            Set the body state to False
        else if section head is not None then
            Append the segment to the section head.
            Set the body state to True.
        else
            Make segment section head
            Set body to False
        end if
    else
    if section head is None then ▷ # for initial segments without section head
            Make segment section head
            Set body to False
        else
            Append the segment to section head.
            Set body to True
        end if
    end if
    if body is False then
        Append section head to complete list of sections
    end if
end for
return Final list of complete sections.
```
